# High treatment success among individuals with rifampicin-resistant tuberculosis in Botswana: A retrospective cohort study

**DOI:** 10.1101/2025.07.14.25331509

**Authors:** Tuelo Mogashoa, Justice T. Ngom, Ontlametse T. Choga, Johannes Loubser, Phenyo Sabone, Tuduetso Molefi, Topo Makhondo, One Stephen, Joseph M. Makhema, Rosemary M. Musonda, Keabetswe Fane, Simani Gaseitsiwe, Rob M. Warren, Sikhulile Moyo, Anzaan Dippenaar, Elizabeth M. Streicher

**Author notes:** These authors contributed equally and share last authorship (Sikhulile Moyo, Anzaan Dippenaar, Elizabeth M. Streicher).

## Abstract

**Background:** The study reports on tuberculosis (TB) treatment outcomes among individuals diagnosed with rifampicin-resistant TB (RR-TB) and assesses predictors associated with treatment outcomes.

**Methods:** We conducted a retrospective study to analyse treatment outcomes of 162 individuals with RR-TB from 2016 to 2023. Treatment outcome proportions were estimated using the binomial exact method with 95% confidence intervals (CI). Predictors of treatment outcomes were assessed using logistic regression models.

**Results:** Of the 162 individuals, 102 (62.7%) were male with a median age of 39 (interquartile range (IQR): 29-50). Most individuals, 78 (48.1%), were from the Greater Gaborone health district, and 88 (54.3%) were people living with HIV (PLWH). Among these individuals, 137 (84.6%, 95% CI [78.2, 89.7]) were successfully treated. Males had higher odds of unfavourable treatment outcomes compared to females (OR = 1.70; 95% CI [0.73, 3.98]). Among those cured, a slightly higher proportion was observed among PLWH (71.8%, 95% CI [62.1, 80.3]) compared to people not living with HIV (PNLWH) (69.2%, 95% CI [58.7, 78.5]). However, the mortality rate was higher in PLWH (10.7%; 95% CI [5.5, 18.3]) compared to PNLWH (6.6%; 95% CI [2.5, 13.8]). Those with a history of TB treatment had 1.03 odds of unfavourable treatment outcomes (95% CI [0.40, 2.73]); however, this association was not statistically significant.

**Conclusion:** Our study shows a high success rate of treatment among individuals with RR- TB, with no significant difference based on sex, TB treatment history, or HIV status. Higher mortality among PLWH highlights the need for targeted interventions among high-risk groups.

## 1. Introduction

Tuberculosis (TB) remains one of the leading causes of death worldwide, despite being curable and preventable. The emergence and spread of Drug-resistant TB (DR-TB), particularly multidrug-resistant (MDR) or rifampicin-resistant (RR) TB, has made TB prevention and treatment more complicated and challenging, posing a significant public health threat. In 2023, the World Health Organisation (WHO) reported an estimated 180,000 laboratory-confirmed MDR/RR-TB cases, with a global treatment success rate of 68% (1) indicating persistent challenges in achieving optimal treatment outcomes. In the WHO African region, approximately 22,500 laboratory-confirmed MDR\RR-TB cases were reported in 2023, with a regional treatment success rate of 73%, which exceeds the WHO target (1, 2). Botswana is listed among the top 30 high TB/HIV burden countries and reports an estimated MDR/RR-TB incidence of 10 per 100,000 (1). However, data on treatment outcomes among people with MDR/RR-TB remain limited. Treatment outcomes serve as critical indicators for assessing the effectiveness of TB prevention and care programs (3).

In Botswana, individuals diagnosed with RR-TB are treated using a standardised treatment regimen following the WHO and national treatment guidelines (4). However, some people may receive individualised treatment based on their previous TB history and drug susceptibility testing (DST) (5). Several clinical, demographic, and socio-economic factors influence treatment outcomes, including HIV infection, malnutrition, smoking, previous TB treatment, and diabetes (6–8). Previous studies have highlighted age, sex, comorbidities, and body mass index (BMI) as key determinants of treatment success (9–11). Understanding the factors associated with treatment outcomes is essential for informing targeted interventions and evidence-based policy development within the National Tuberculosis Program. Although several studies in Botswana have assessed TB treatment outcomes, few have focused specifically on individuals with RR-TB (12–14). This study aims to characterise treatment outcomes among individuals diagnosed with RR-TB in Botswana and to identify predictors associated with these outcomes.

## 2. Materials and Methods

### 2.1 Study design and population

This study employed a retrospective cohort design to evaluate treatment outcomes among individuals diagnosed with RR-TB in Botswana between 1 January 2016 and 30 June 2023. The study population consisted of individuals with laboratory-confirmed RR-TB who were enrolled in treatment programs consistent with National TB Treatment guidelines (8) at designated TB treatment clinics in Botswana. Data were extracted from national and district TB surveillance databases, including demographic characteristics, clinical history, microbiological test results, and treatment outcomes.

Eligible participants included individuals aged 18 years or older with complete treatment records. The primary outcome was treatment success (cured) versus unfavourable outcomes (treatment failure, loss to follow-up [LFU], or death). Key predictor variables included previous TB treatment history, sex, HIV status, smear results, health district, and *Mycobacterium tuberculosis* (*M.tb)* lineages.

### 2.2 Phenotypic drug susceptibility testing, DNA extraction, whole genome sequencing and bioinformatic analyses

Phenotypic susceptibility testing, DNA extraction, whole-genome sequencing, and bioinformatic analyses were performed as previously described (15, 16). Drug resistance genotypes for both first- and second-line antibiotics and lineages were inferred using TB-Profiler software, version 4.4.2 (17, 18).

### 2.3 Treatment outcomes and definition

Treatment outcomes were assigned based on patient progress, following the WHO guidelines (19). The treatment outcomes include cured, treatment failure, died, and LFU. Definitions are presented below in Table 1:

**Table 1.** TB treatment outcomes and definitions.

| Treatment outcome | Definition |
| --- | --- |
| Cured | A pulmonary TB patient with bacteriologically confirmed TB at the beginning of treatment who completed treatment as recommended by the national policy with evidence of bacteriological response and no evidence of failure. |
| Died | A patient who died before starting treatment or during the course of treatment. |
| Treatment failure: | A patient whose treatment regimen needed to be terminated or permanently changed to a new regimen or treatment strategy. |
| LFU | A TB patient who did not start treatment or whose treatment was interrupted for two consecutive months or more. |

In this study, unfavourable treatment outcomes included LFU, treatment failure, and death, while favourable treatment outcomes included those who were cured.

### 2.4 Statistical analysis

Baseline demographics were summarised by frequencies and percentages for categorical variables (with 95% confidence intervals, [CI]) and medians with interquartile range (IQR) for continuous variables. The proportions of treatment outcomes were estimated with a 95% CI using the binomial exact method. Predictive factors associated with unfavourable treatment outcomes were assessed using a univariate logistic regression (Firth’s logistic regression). Firth’s logistic regression was used to reduce the bias in maximum likelihood estimates of the coefficients (20). The use of Firth’s logistic method helps reduce bias when there is a strong imbalance in the outcome, as is the case with our outcome variable. The variables evaluated were age, sex, HIV status, site of TB disease, previous treatment history, and *M.tb* lineage; p-values less than 0.05 indicated statistical significance. All statistical analyses were performed in STATA version 18 (Stata Corp, College Station, TX, USA) (21)

## 3. Results

A total of 202 individuals diagnosed with RR/MDR-TB were identified in Botswana between 1 January 2016 and 30 June 2023 and initiated on treatment. Of these, 40 individuals were excluded because they did not meet the study inclusion criteria. Additionally, 36 were not evaluated for treatment outcomes, while four were still undergoing treatment. Those with missing treatment outcome data were also excluded (**Figure 1**). 102 (63%) were male with a median age of 39 (IQR, 29,50). The largest proportion of individuals were from the Greater Gaborone health district, 78 (48.1%); 88 (54.3%) were people living with HIV, while 1 (0.6%) had an unknown HIV status. All individuals with a confirmed HIV diagnosis at the time of TB diagnosis were initiated on antiretroviral treatment (ART) and cotrimoxazole preventive therapy (CPT). Most individuals were newly diagnosed with RR-TB (121, 74.7%) (**Table 2**).

**Figure 1.**
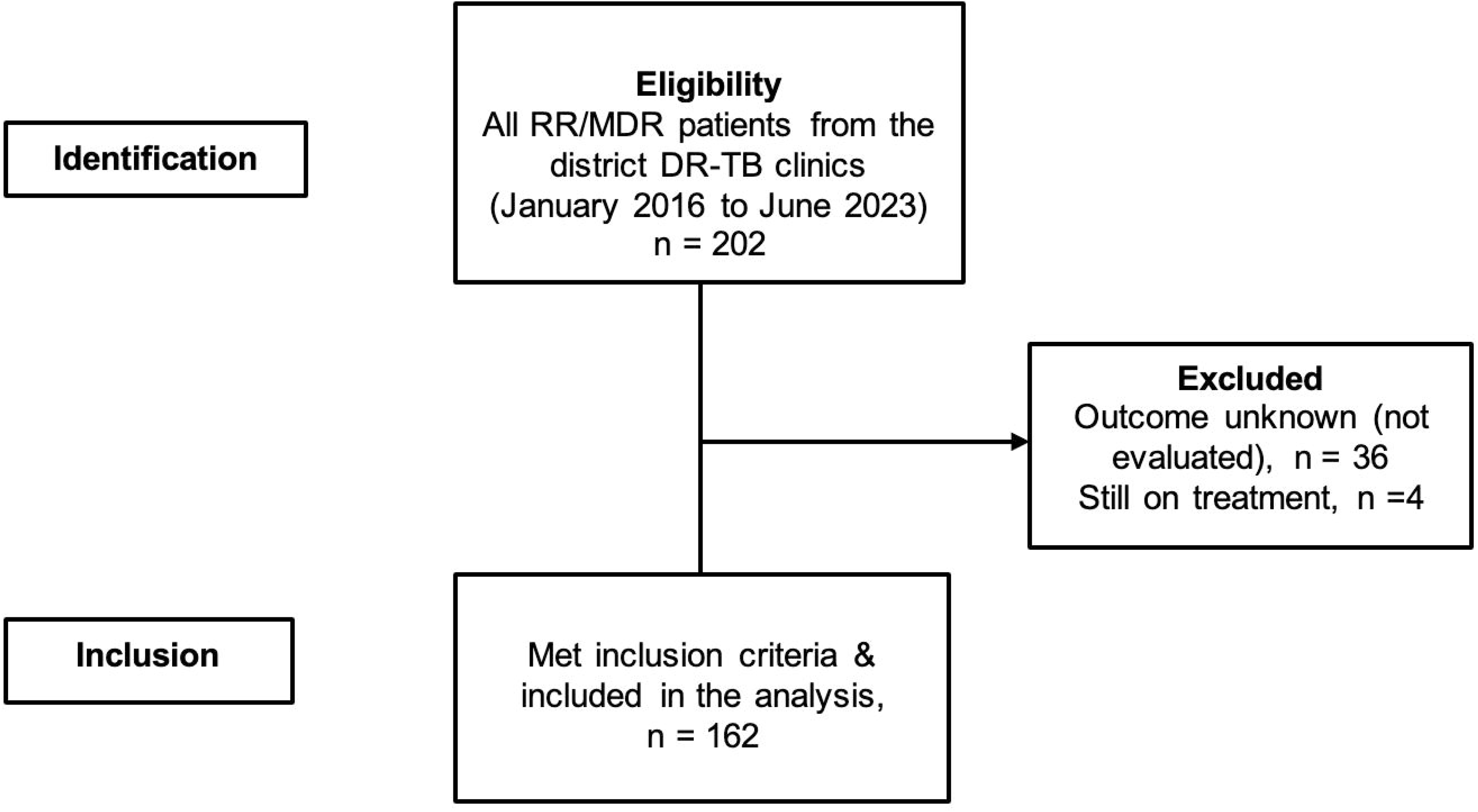
Participant identification and inclusion criteria

**Table 2:** Demographic and clinical characteristics of 162 individuals with rifampicin-resistant tuberculosis.

| Characteristic | N | % |
| --- | --- | --- |
| **Sex (N=162)** |  |  |
| Female | 60 | 37.0 |
| Male | 102 | 63.0 |
| **Median Age in years (Q1, Q3)** | **39 (29, 50)** |  |
| **Health district** |  |  |
| Greater Gaborone | 78 | 48.1 |
| Greater Francistown | 31 | 19.1 |
| Greater Palapye | 21 | 13.0 |
| Greater Maun | 22 | 13.6 |
| Other* | 10 | 6.2 |
| **HIV status** |  |  |
| Negative | 73 | 45.1 |
| Positive | 88 | 54.3 |
| Unknown | 1 | 0.6 |
| **Smear results** |  |  |
| Negative | 36 | 22.2 |
| Positive | 126 | 77.8 |
| **Site of TB disease** |  |  |
| Pulmonary | 158 | 97.5 |
| Extra-pulmonary | 4 | 2.5 |
| **Previous treatment history** |  |  |
| New | 121 | 74.7 |
| Previously treated | 41 | 25.3 |
| ***M.tb* lineages** |  |  |
| Lineage 4 | 98 | 60.5 |
| Other lineages<sup>#</sup> | 64 | 39.5 |
| **Genomic DR-TB type** |  |  |
| RR-TB | 32 | 19.8 |
| MDR-TB | 99 | 61.1 |
| Pre-XDR-TB | 27 | 16.7 |
| Sensitive | 4 | 2.5 |

*Other\* included Ghanzi (n=5), Kgalega (n=4) and Boteti (n=1) and health districts;* *observations less 10 per health district; HIV- human immunodeficiency virus, other lineages#* *includes lineage 1 (n=36), lineage 2 (n=24), lineage 3 (n=2) and mixed lineages (n=2)*

### 3.1 Treatment outcomes

Of the 162 individuals with complete TB treatment outcomes, 137 (84.6%, 95% CI [78.2, 89.7]) were successfully treated; 18 (11.1%, 95% CI [6.7, 17.0]) died during treatment, while 2 (1.2%, 95% CI [0.2, 4.3]) had treatment failure. Additionally, five individuals (3.1%, 95% CI [1.0, 7.1]) were lost to follow-up (**Table 3**).

**Table 3:**
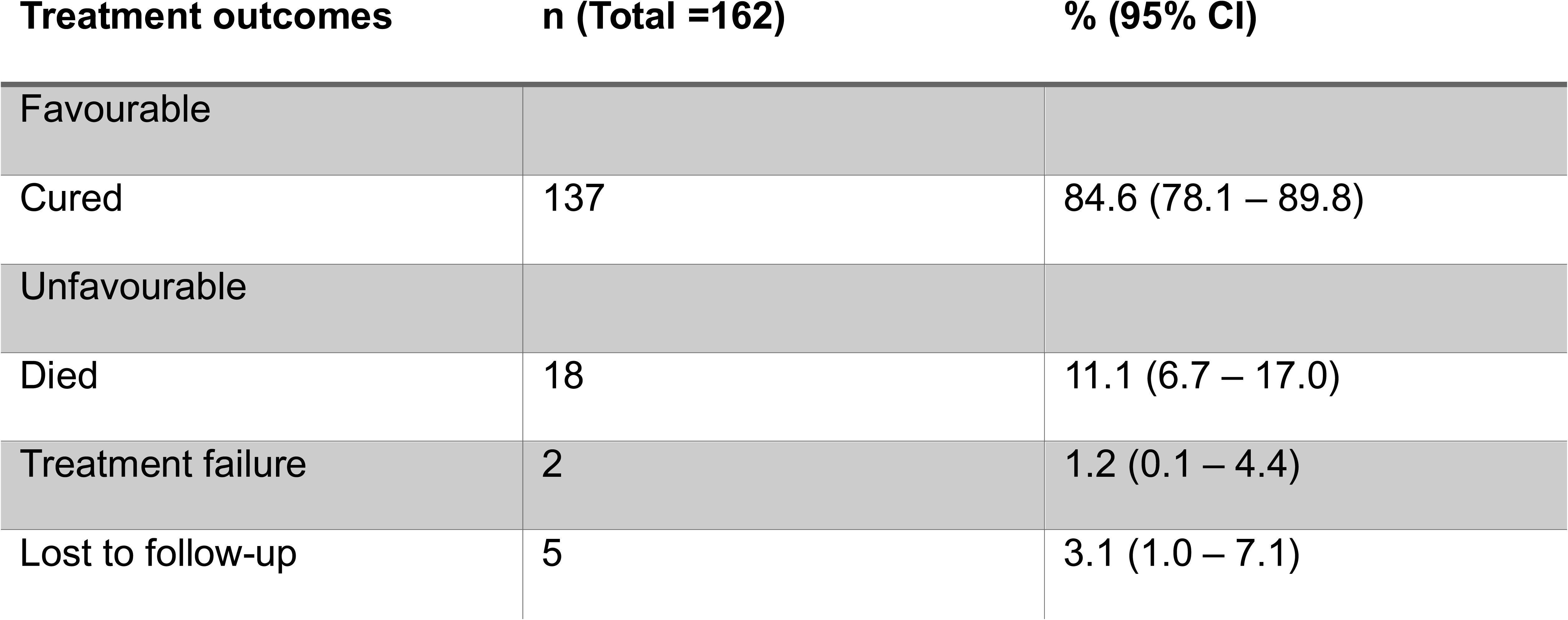
Tuberculosis treatment outcomes of 162 individuals based on WHO classification.

| Treatment outcomes | n (Total =162) | % (95% CI) |
| --- | --- | --- |
| Favourable |  |  |
| Cured | 137 | 84.6 (78.1 – 89.8) |
| Unfavourable |  |  |
| Died | 18 | 11.1 (6.7 – 17.0) |
| Treatment failure | 2 | 1.2 (0.1 – 4.4) |
| Lost to follow-up | 5 | 3.1 (1.0 – 7.1) |

Figure 2 presents a comparison of TB treatment outcomes between people not living with HIV (PNLWH) and people living with HIV (PLWH). Among those who were cured, a higher proportion was observed among PLWH (71.8; 95% CI [62.1, 80.3]) compared to PNLWH (69.2, 95% CI [58.7, 78.5]). However, the proportion of deaths was higher in PLWH (10.7; 95% CI [5.5, 18.3]) compared to PNLWH (6.6; 95% CI [2.5, 13.8]). Conversely, the proportion of individuals who were lost to follow-up was lower in PLWH (1.9; 95% CI [0.2 - 6.8]) compared to PNLWH (2.2; 95% CI 0.3 - 7.7). Treatment failure was observed in 2.2% of PNLWH (Figure 2).

**Figure 2:**
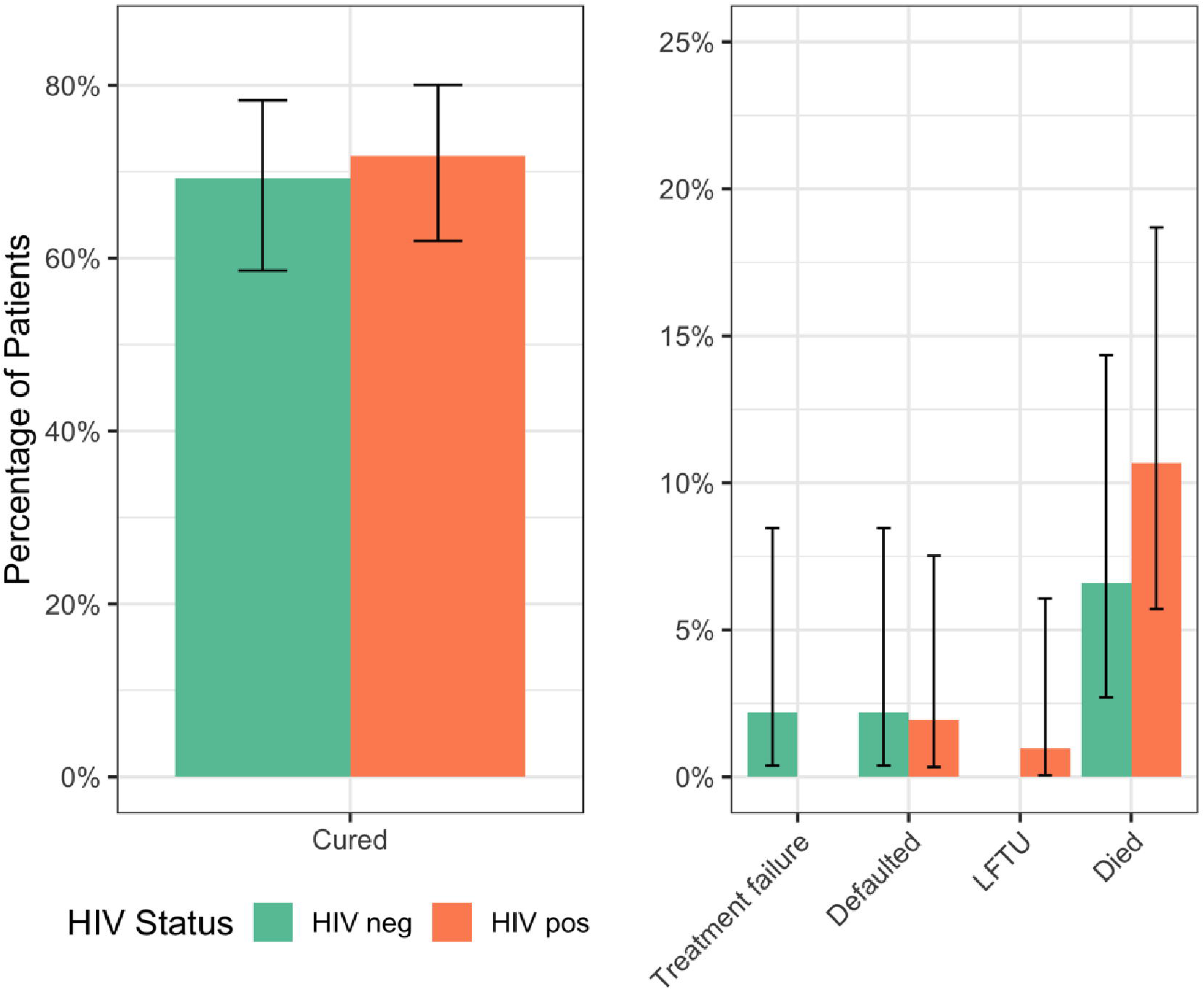
Tuberculosis treatment outcomes of individuals with rifampicin-resistant tuberculosis stratified by HIV status

Males had 1.70 times the odds of unfavourable treatment outcomes compared to females and the differences between the groups were not statistically significant (OR = 1.70; 95% (95% CI [0.73 - 3.98]) The median age was 39 (Q1, Q3: 29, 50), and individuals aged below 39 had 1.20 (95% CI [0.63, 2.30]) odds of an unfavourable treatment outcome compared to older individuals. All people living with HIV (PLWH) were on antiretroviral therapy (ART) and cotrimoxazole preventive therapy (CPT), PLWH showed slightly lower, but non-significant odds of unfavourable outcomes compared to people not living with HIV (OR = 1.18, 95% CI 0.50 - 2.79). Additionally, individuals with a prior history of TB treatment had 1.03 (95% CI [0.40 - 2.73]) odds of unfavourable treatment outcomes. However, none of these associations were statistically significant (**Table 4**).

**Table 4:**
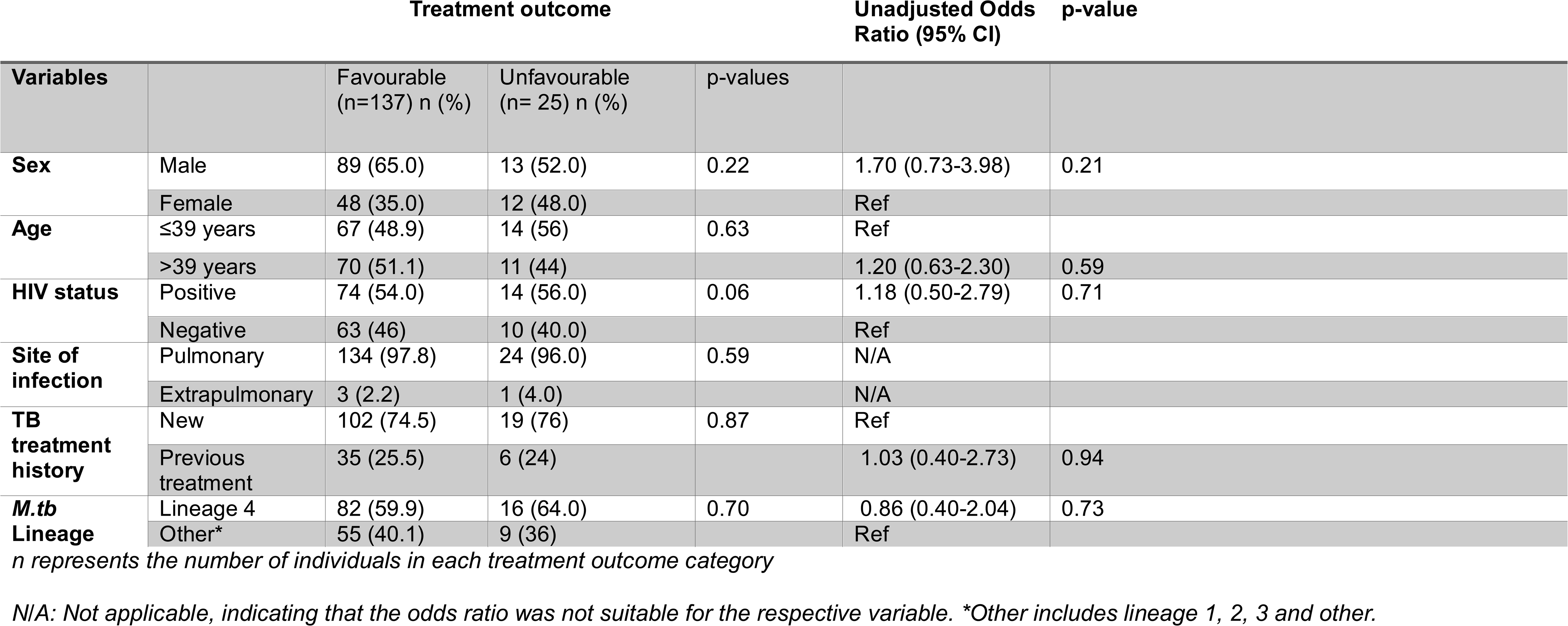
Predictive factors of unfavourable treatment outcomes (n=162)

## 4. Discussion

This study examined the influence of various clinical and demographic factors on TB treatment outcomes, yielding an 84.6% treatment success rate. The high treatment success rate highlights the effectiveness of the treatment regimen used in this cohort. This is similar to what has been previously reported (22–24) and higher relative to the global RR-TB treatment success rate reported to the WHO (1). The high success rate may likely be attributed to advances in TB diagnosis and several other interventions, such as direct observed therapy (DOTS) and the use of molecular diagnostic tools, including the Cepheid Gene Xpert MTB/RIF system and line probe assays (25), which have made it possible to detect RR-TB earlier. Additionally, the advances in HIV diagnosis and expanded ART access in Botswana may have contributed to the treatment success rates among PLWH (22). The proportion of PLWH co-infected with TB achieving treatment success was similar to treatment success rates among PNLWH. Similar findings have been previously reported in other studies (26, 27).

In contrast to the high treatment success observed in this cohort, a similar study in Zimbabwe and Colombia reported high unfavourable treatment outcomes among individuals with RR-TB (11, 28). This highlights the significance of context-specific factors, including healthcare infrastructure, adherence support, and socioeconomic conditions, in influencing treatment outcomes. However, the notable proportion of unfavourable treatment outcomes, particularly deaths in this study, underscores the need for closer patient monitoring and targeted interventions to improve adherence and survival.

The low rates of unfavourable treatment outcomes were coupled with challenges in identifying predictors of these outcomes. Our findings show that none of the evaluated patient characteristics were significantly associated with unfavourable treatment outcomes. Being male, aged less than 39 years, had increased odds of experiencing treatment failure; however, the associations were not statistically significant. These findings differ from what has been previously reported in other studies (29), where being male was statistically associated with unfavourable treatment outcomes. Similarly, HIV status was not statistically associated with treatment success, although a higher mortality was observed among PLWH compared to PNLWH. In this cohort, factors traditionally considered predictors of unfavourable treatment outcomes, such as age, HIV status, and sex, did not show statistically significant associations; this differs from findings in previous studies (8, 27, 30). Prior research has demonstrated that socioeconomic factors such as educational level and type of employment, and behavioural factors, such as smoking and alcohol use, substantially affect treatment outcomes in patients with RR-TB (7). However, in our study, these variables were either not collected or insufficiently documented, making them unavailable for analysis.

Simplifying treatment outcome definitions in programmatic settings may be necessary to improve accuracy and consistency in the reporting of outcomes. A previous study suggested that simplifying WHO treatment outcome definitions may better account for variations in sputum culture timing and the definition of conversion in programmatic settings (31). The study also demonstrated that simplified definitions were more effective in identifying individuals who are at a higher risk of treatment failure than the WHO-based criteria (31). This highlights the potential benefits of adapting treatment outcome definitions and classifications to enhance patient monitoring and improve TB management strategies.

We acknowledge that our study has several limitations that may affect the interpretation and generalizability of our findings. The data were obtained retrospectively from TB registers; therefore, it was not possible to access crucial patient-level information such as comorbidities (e.g., diabetes, CD4 and viral load counts [for PLWH], BMI, smoking habit, education level, alcohol consumption, and substance use). The absence of these variables could have influenced our analyses, potentially introduced residual confounding and limited our ability to fully adjust for factors known to affect TB treatment outcomes negatively. Socioeconomic factors such as level of education and employment status, as well as behavioural factors like smoking status and alcohol consumption, have been shown in other studies to have an impact on patient TB treatment outcomes (7, 8). However, in our study, these variables were either not collected or insufficiently documented, making them unavailable for analysis. Our study may have been underpowered to detect statistically significant differences across the groups. Many comparisons yielded broad and overlapping confidence intervals, increasing the possibility of a Type II error. Limited statistical power may have prevented the detection of true associations, and the absence of statistically significant associations should not be interpreted as evidence of no effect. Future prospective studies with richer patient-level data and larger sample sizes are warranted to validate these results.

We were unable to evaluate the effectiveness of each individual’s regimen due to the unavailability of detailed treatment histories, including drug doses, treatment modifications, adverse events, adherence measures, and reasons for regimen changes. Treatment heterogeneity as well as the use of newer TB drugs such as bedaquiline could significantly influence treatment outcomes; however, these data were not available, and their impact on treatment outcomes was not measured. We acknowledge that the absence of these data may have affected our findings and could partly explain the variability observed in this study. This limitation also restricted our ability to assess regimen-specific outcomes such as the comparative effectiveness of different drug combinations or the role of newer drugs in improving treatment outcomes. Future studies should incorporate data such as detailed treatment and adherence data to accurately assess the impact of regimen-specific factors on MDR/RR-TB treatment outcomes. Despite these limitations, the study provides valuable insights based on real-world programmatic data, and the findings remain relevant for informing TB control efforts in similar settings where comprehensive data collection may not always be feasible.

In conclusion, the high treatment success rate of 84.6% reflects a positive impact of improved diagnostics and treatment strategies in Botswana. Although no statistically significant associations were found between clinical and demographic factors and unfavourable treatment outcomes among individuals with MDR/RR-TB, a higher mortality in PLWH may have some clinical relevance. Enhanced patient monitoring and targeted interventions, particularly for vulnerable groups like PLWH, remain essential to improve treatment outcomes further. Future studies with a larger sample size, more comprehensive clinical and socioeconomic data are warranted to explore these factors further, confirm the trends observed in our research, and better identify predictors of unfavourable treatment outcomes.

## Institutional Review Board statement

The study was approved by the Stellenbosch University Health Research Ethics Committee (HREC) [HREC Reference No: S22/12/271 (PhD)] and the Ministry of Health Human Research Development Division [Reference No: HPRD: 6/14/1]. Permission was granted to use de-identified participant clinical information; therefore, informed consent was waived. This study was conducted following the guidelines and regulations established by the ethics committees of the respective institutions and the Declaration of Helsinki.

## Data availability statement

The original contributions presented in the study are publicly available. Sequencing data used in this study have been deposited in the European Nucleotide Archive (ENA) under project accession number PRJEB83872.

## Acknowledgements

We want to thank the Ministry of Health, the National Tuberculosis Reference Laboratory, the National Health Laboratory, the Botswana National Tuberculosis Program and Victus Global Botswana for their support and collaboration in this project.

## Author contribution

Conceptualization: TM, RMW, EMS Methodology: EMS, RMW, AD, SM, TM, JHL, JTN Data curation and formal analysis: TM, OTC, JHL SM, EMS, JTN, PS Writing original draft: TM, SM, JTN Writing, review & editing: TM, OTC, EMS, RMW, AD, JHL, JTN, SG, SM, CM, OS, PS, TMM, KF, RMW, JM; Supervision: EMS, RMW, AD, SM, SG; Project administration: EMS, TM, SM, RMW; Funding acquisition: TM, SM, EMS. All authors read and approved the final manuscript.

